# Integrating the cognitive sequelae after temporal lobe surgery into daily life: a mixed methods approach to the consequences of average to severe memory decline

**DOI:** 10.64898/2026.08.03.26359089

**Authors:** J. Taube, D. Middendorf, G. Taube, E. Francke, C. Reinecke, L. Helmstaedter, J.-A. Witt, V. Borger, A. Racz, R. Surges, C Helmstaedter

**Affiliations:** Department of Epileptology, University Hospital Bonn, Bonn, Germany; Clinic for Geriatric Psychiatry and Cognitive Disorders, University Hospital Bonn, Bonn, Germany; Fachhochschule des Mittelstands GmbH - University of Applied Sciences, Bielefeld, Germany; Behavioral Neurology Research Group, Department of Neurology, University Hospital Knappschaftskrankenhaus Bochum, Ruhr University Bochum, Bochum, Germany; Department of Neurosurgery, University Hospital Bonn, Bonn, Germany

**Keywords:** Epilepsy, Temporal Lobe, Epilepsy Surgery, Memory Disorders, Independence in Activities of Daily Living, Mixed Methods Research

## Abstract

**Background:** Temporal lobe epilepsy surgery (TLS) is an effective treatment for drug-resistant focal epilepsy but is often associated with cognitive decline. A key unresolved question is how to distinguish average from severe memory loss and how these levels differentially affect everyday life. We addressed this question applying patient-derived norms of memory decline and conducting qualitative interviews with patients experiencing expected or unexpectedly severe memory loss.

**Methods:** Regression-based normative change criteria for postoperative verbal memory loss were derived from a single-center cohort of 806 patients. Two matched groups of four patients each were selected from the severe memory decline (SMD; below the 5th percentile) and average memory decline (AMD; 20^th^ – 75^th^ percentile) ranges. In-depth, semi-structured narrative interviews were analyzed using qualitative content analysis with a combined deductive-inductive approach.

**Results:** AMD narratives emphasized recovery, with surgery integrated into a continuing sense of self. In contrast, SMD descriptions focused on persistent symptoms, continued treatment, and illness despite meaningful seizure reduction. Patients with SMD also reported limited information, insufficient psychological preparation or postoperative cognitive rehabilitation.

**Conclusions:** Whereas AMD was generally manageable and successfully integrated into everyday life, SMD disrupted identity, autonomy, and expected life trajectories. Current surgical pathways appear to address these cognitive sequelae insufficiently. Improved expectation management, together with structured preoperative counseling and postoperative rehabilitation, may facilitate adaption to memory decline and improve long-term functioning and quality of life after surgery.

**What is already known on this topic:** Temporal lobe epilepsy surgery (TLS) is an effective treatment for drug-resistant epilepsy but is frequently associated with postoperative memory decline. Although the cognitive risks of surgery are well established, it remains unclear when memory decline becomes a clinically meaningful complication and how different degrees of decline affect patients’ everyday lives.

**What this study adds:** While patients have traditionally been counselled primarily about the probability of experiencing memory decline after TLS, our findings shift the focus towards the severity and consequences of such decline. If memory loss occurs, the average magnitude of decline appears to be largely manageable in everyday life, whereas disabling memory impairment should be considered a very rare complication.

**How this study might affect research, practice or policy:** These findings support integrating individualized expectation management, structured neuropsychological follow-up, and cognitive and psychological pre-/rehabilitation into routine epilepsy surgery care, particularly for patients at high risk of severe memory decline.

## Introduction

Temporal lobe epilepsy surgery (TLS) offers substantial improvements in seizure control [1,2], and is often viewed as a means of restoring quality of life (QOL) by reducing epilepsy’s impact on independence, social participation, employment, and everyday functioning [3,4]. Hence, postoperative QOL is not determined by seizure control alone, but is also strongly shaped by cognitive outcomes, particularly memory.

Postoperative cognitive decline, especially in verbal memory, is a well-documented consequence following TLS [5–8]. However, determining when postoperative change constitutes a clinically meaningful cognitive complication remains challenging. Contemporary epilepsy surgery is increasingly tailored to the individual, with surgical approach, extent of resection, underlying pathology, and functional anatomy varying substantially between patients. These advances maximize seizure control while minimizing collateral damage, yet different postoperative memory changes can be expected, making it difficult to define a threshold for clinically significant cognitive loss across surgical procedures [9,10].

Standard frameworks for surgical complications typically capture events such as cerebrospinal fluid leak, intracranial or extracranial infection, or focal neurological deficits [11], but fail to capture cognitive difficulties that substantially affect daily functioning despite not meeting criteria for classical adverse events. This limitation is particularly relevant for memory impairments, which for some patients outweigh the benefits of successful seizure control and largely impact postoperative QOL [12]. Identifying when postoperative memory decline represents an expected consequence versus a clinically significant complication remains an important but unresolved challenge.

Most previous studies rely on group-level or average change scores, which do not account for individuals whose postoperative trajectory falls outside the expected course. Although meta-analytic evidence indicates overall improvements in QOL following epilepsy surgery, these gains depend strongly on favorable seizure outcome and preserved cognitive functioning. Moreover, average improvements mask considerable differences between individuals. [13,14]. Consequently, quantitative outcome measures provide limited insight into how patients experience and adapt to cognitive changes that exceed the expected postoperative course.

The present study addresses this gap by combining an individualized definition of cognitive outcome with qualitative investigation of patients lived experiences. Using standardized regression-based change norms derived from a large surgical cohort, we classified postoperative memory performance according to whether cognitive change fell within or beyond the expected range for an individual patient. This approach enables the identification of patients with severe postoperative memory decline and those showing changes in the expected range. By integrating individual-level neuropsychological classification with qualitative analysis of lived experience, we investigate how these groups differ in their subjective evaluation of postoperative daily-life functioning.

## Methods

### Study Design

We employed a qualitative comparative design embedded within a mixed-methods framework. Regression-based normative change criteria derived from a large single-center epilepsy surgery cohort with temporal lobe epilepsy (n = 806 allowed classification of patients into two groups (severe vs average loss) using pre- to postoperative verbal memory loss data (in percentiles). The qualitative component comprised in-depth, semi-structured interviews, which were analysed using qualitative content analysis in MAXQDA 22. Interview transcripts were systematically coded, paraphrased, condensed, and organised into thematic categories.

Integration of the quantitative and qualitative components occurred at two stages: first, during participant selection, where individuals were grouped; and second, at the interpretive synthesis stage, where category-based findings were compared across the cognitively defined groups to identify systematic differences in illness narratives, pre-operative expectations, and evaluations of postoperative daily-life functioning.

The study was conducted in accordance with the Declaration of Helsinki. Ethical approval was obtained from the University Hospital Bonn review board prior to data collection. All participants provided written informed consent.

### Participants

All participants had temporal lobe epilepsy (TLE) and were recruited during epilepsy surgery follow-up at a university epilepsy centre in Germany. Inclusion criteria were: (1) diagnosis of drug-resistant TLE; (2) having undergone resective epilepsy surgery at least twelve months prior to the interview; (3) having completed comprehensive pre-operative and postoperative memory assessment; (4) being 18 years or older at time of surgery; and (5) sufficient German language proficiency. Exclusion criteria were: incomplete memory data and surgery less than twelve months prior to the interview.

### Standardized change norms

To classify verbal memory decline, we calculated age-dependent regression-based norms from a reference cohort (n = 806). Participants had a mean age of 29.75 years (SD=12.95) and seizure onset age of 12.57 years (SD=10.08); 50.4% were female. Left- and right-sided surgeries were equally represented (left:right ratio=1.04). Surgical procedures are detailed in Table 1.

**Table 1.** Types of Surgery for Reference Cohort.

| Surgery Type | Percentage |
| --- | --- |
| Amygdalohippocampectomy | 33 |
| Amygdalohippocampectomy + lobectomy | 23 |
| Lesionectomy | 16 |
| Partial resection | 7 |
| Partial resection + amygdalohippocampectomy | 7 |
| Other | 14 |

Participants were classified based on the free recall subcomponent of the “Verbaler Lern- und Merkfähigkeitstest” (VLMT), the German version of the Rey Auditory Verbal Learning Test (VLMT) [15]. Delayed recall was selected as the primary index. reflecting consolidation and retention of newly learned verbal material. This measure is most closely connected to temporomesial pathology, and sensitive to TLS, particularly on the left side [16]. Additionally, it has proven to be an ecologically valid measurement tool of verbal learning [17–21]. Compared to immediate recall or learning-trial totals, delayed recall more specifically captures hippocampal-dependent long-term memory function and shows the largest and most reliable postoperative change in TLS cohorts.

Regression-based norms were computed using general additive models for location, scale, and shape (GAMLSS), accounting for potential non-linear age effects and age-dependent changes in variability. The Box-Cox Power Exponential (BCPE) distribution accommodated deviations from normality. The final model included a second-order polynomial for age in the location parameter and a linear age term in the dispersion parameter. This framework enabled derivation of age adjusted expected values and age-specific variability estimates for individual-level standardization [22–24]:

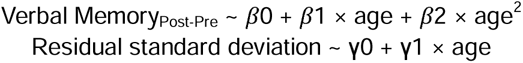

Patients with the greatest pre- to postsurgical decline (SMD group, below 5th percentile) were contrasted against those with average memory decline (AMD group, 20th–75th percentile). The SMD threshold corresponds to the conventional neuropsychological cut-off for atypical performance. The AMD comparison group represented the expected postoperative trajectory. Eight participants were included: four in the SMD group and four in the AMD group. The lower bound was deliberately set above the SMD threshold to establish a clear separation zone between groups and to exclude borderline cases in the 5th–20th band that would blur the contrast. Similiarly the upper bound was set below the top quartile to exclude atypically favourable outcomes (i.e. exceptional preservation or improvement) that would otherwise confound a decline-focused comparison. This purposive, contrast-maximising sampling strategy prioritised between-group distinctness over statistical representativeness, consistent with the study’s exploratory, hypothesis-generating aims. As shown in **Fig. 1**, the regression-based norm of verbal memory loss is presented with percentile curves. **Tables 2a** and **2b** present participant and clinical characteristics.

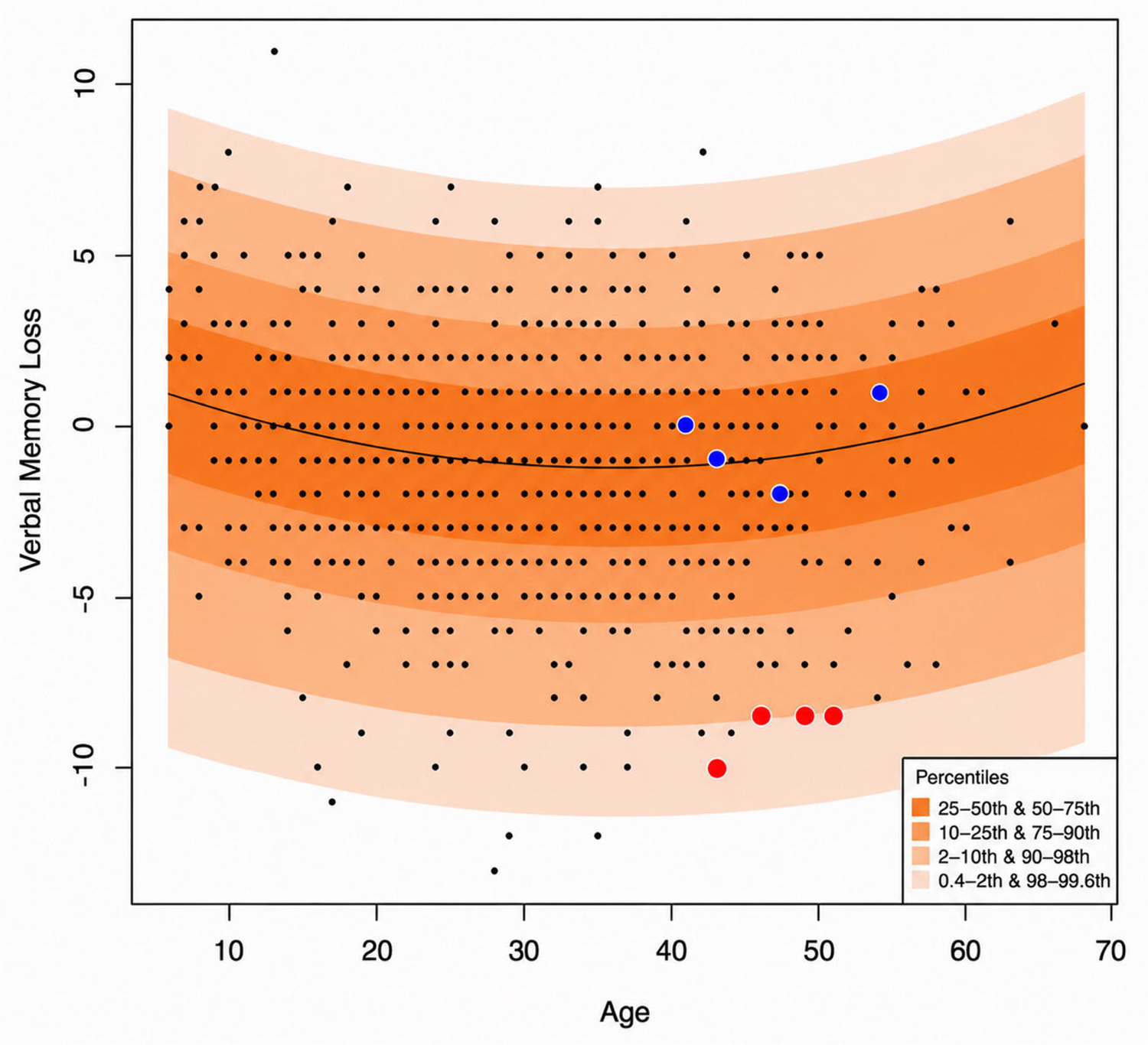

**Table 2a.** Participant Demographic and Clinical Characteristics.

| Group | Pct | ID | Sex | Onset,<br>y | Surgery,<br>y | Interview,<br>y | Side | Procedure | IA | Pathology |
| --- | --- | --- | --- | --- | --- | --- | --- | --- | --- | --- |
| SMD | 03 | 01 | M | 26-30 | 41-45 | 41-45 | Le | ATL + AH | Bitemporal | No HS /<br>gliosis only |
| SMD | 02 | 02 | M | 46-50 | 51-55 | 51-55 | Le | AH | Bitemporal | HS |
| SMD | 03 | 03 | F | 06-10 | 41-45 | 46-50 | Le | SAH | None | HS |
| SMD | 01 | 05 | M | 16-20 | 36-40 | 46-50 | Le | ATL + AH | None | No HS /<br>gliosis only |
| AMD | 62 | 06 | F | 01-05 | 36-40 | 31-35 | Le | SAH | Left temporal<br>+ occipital | HS |
| AMD | 21 | 09 | F | 26-30 | 36-40 | 41-45 | Le | SAH | Bitemporal | No HS /<br>gliosis only |
| AMD | 23 | 10 | M | 06-10 | 51-55 | 66-70 | Ri | SAH | Bitemporal | HS |
| AMD | 23 | 25 | F | 01-05 | 41-45 | 46-50 | Ri | SAH | None | HS |

*Abbreviations.* AH, amygdalohippocampectomy; AMD, average memory decline; ATL, anterior temporal lobectomy; F, female; HS, hippocampal sclerosis; IA, invasive assessment; M, male; PCT, percentile of loss; SAH, selective amygdalohippocampectomy; SMD, severe memory decline; Y, age-range in years;

**Table 2b.** Seizure history, treatment.

| ID | IA complications | Pre focal | Pre GTCS/ FBTC | Post focal | Post GTCS/ FBTC | Pre ASM | Post ASM | Periop course | Comorbidities |
| --- | --- | --- | --- | --- | --- | --- | --- | --- | --- |
| 01 | subarachnoid hemorrhage, right temporal; ICB, left temporal | 10/month | None since 2019 | — | 1 at 13 months postop | 2 | 2 | Postop hemorrhage, expressive aphasia, right upper quadrantanopia | HSV encephalitis; HIV; Hodgkin disease treated with chemotherapy; prior substance abuse |
| 02 | Moderate expressive aphasia; mild receptive aphasia | 4/month | 1 | — | 2 within 6 months | 2 | 2 | Postop subdural hematoma; mild expressive and receptive aphasia | — |
| 03 | — | 2/month | — | 4/month | None | 2 | 2 | Immediate postop aphasia and word-finding difficulty | Depression; obesity |
| 05 | — | 8/month | — | 1/month | None | 3 | 3 | Expressive language disturbance | Clival chordoma treated with proton therapy and Gamma Knife |
| 06 | — | 2/month | — | 1/month | None | 4 | 4 | Status epilepticus in February 2024 | — |
| 09 | — | 3/month | 1 | — | None | 2 | 1 | Emotional/self-perception difficulties | Prior substance abuse |
| 10 | — | 10/month | 4 | — | None | 3 | 3 | Elevated depressive symptoms | — |
| 25 | — | 30/month | — | 2/month | None | 2 | 2 | — | Hydrocephalus following early childhood meningitis with VP shunt |

### Data Collection

Semi-structured narrative interviews were conducted by trained interviewers in person or hybrid online. The interview guide was developed iteratively in consultation with the clinical team. Participants were invited first to tell the story of their experiences with epilepsy, including the period leading to the surgical decision, the surgical experience itself, and life since surgery. Structured probing questions followed regarding occupational functioning, social relationships, identity and self-perception. Interviews were recorded and transcribed verbatim. Interview duration ranged from approximately 60 to 90 minutes.

### Data Analysis

Qualitative content analysis was conducted in MAXQDA (v22) using a deductive-inductive approach. An initial code frame was developed a priori based on the interview guide and epilepsy surgery psychosocial outcome literature. In a second cycle, inductive open coding was conducted by an experienced social scientist (>19 years), blind to group membership, to identify additional themes. Coding followed a systematic category-based procedure, with all coded segments reviewed within each category to assess consistency and differentiate subthemes and between-group patterns. To enable group comparisons while accounting for differences in transcript length, coded segments were quantified as within-participant proportions of total coded material.

### Reflexivity and Quality Criteria

The research team consisted of clinical neuropsychologists and a qualitative methodologist with expertise in epilepsy surgery outcomes. Clinical familiarity was both an analytic resource and a potential source of bias, addressed through reflexive documentation and blinded secondary coding. Credibility was supported by prolonged data engagement, member checking with two participants, and negative case analysis. Transferability is limited by the single-centre, German-language sample; findings are intended as hypothesis-generating.

We used the STROBE reporting guideline(1) to draft this manuscript, and the STROBE reporting checklist(2) when editing, included in supplement A.

## Results

### Improvement in AMD vs Symptom and Illness Focus in SMD

When code frequencies were expressed as the proportion of all coded segments within each group, several differences emerged. Perceived improvement after surgery appeared more prominent in the AMD than in the SMD group (17.2% vs 3.5%), even though the AMD group was more often ambivalent in the evaluation (7.2% vs 5.0%). The SMD group tended to talk more about new or changed symptoms, medication and ongoing treatment (8.2% vs 2.6% in AMD) after surgery (11.3% vs 4.1% in AMD). Notably, the SMD group more often emphasized preoperative illness history/epilepsy origin (6.3% vs 2.6%) and acceptance/illness framing (4.1% vs 0.5%). The AMD group tended to show greater emphasis on realistic expectations regarding surgery (4.4% vs 0.4% in SMD). Overall, AMD accounts were more strongly characterised by improvement-oriented evaluations of outcome, whereas SMD accounts placed greater emphasis on ongoing symptoms, treatment burden, and illness context. **Figure 2** shows all frequencies across all codes. Sensitivity analysis recomputing group code frequencies after excluding the AMD case at the 62nd percentile did not alter the direction of any of the principal between-group differences.

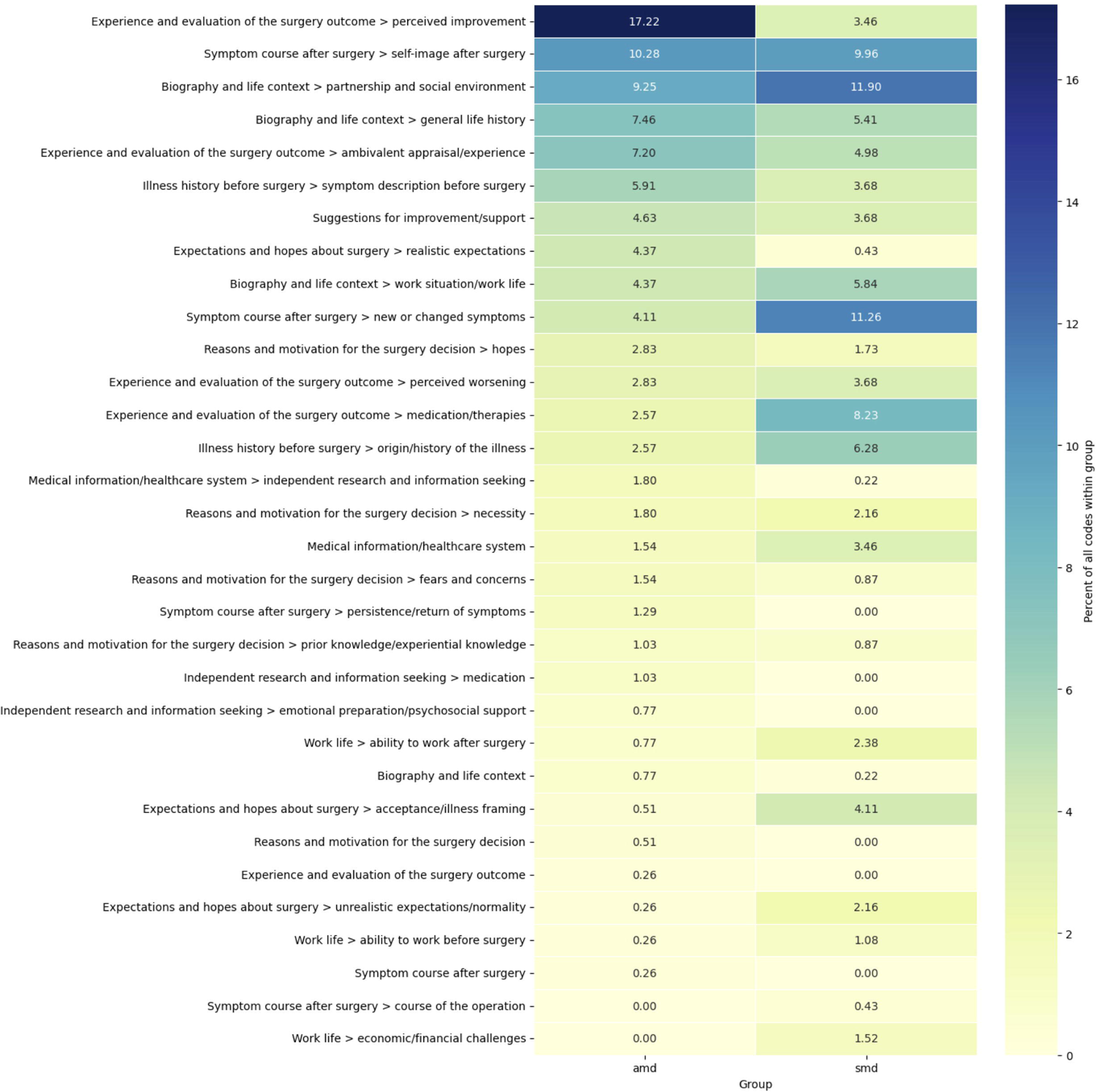

### Pre-operative framing and expectations differ substantially between groups

Participants with SMD described epilepsy as a prolonged and destabilising biographical crisis marked by loss of control, failed medication attempts, and therapeutic exhaustion (01: „ […] and it wasn’t until the medications started to work less and less - and I think I’ve tried every medication out there by now - that I finally decided to seek treatment.”) Surgery was commonly framed not as an active choice, but as the absence of alternatives (05: „[…] since we’d tried everything medication-wise and there were no other options left, and since I, um, had been having extreme seizures lately and was just, um, lying in bed all the time, they told me, um, as advice, that there were almost no other options left […]”). One participant recalled being told there was a two-thirds chance of improvement but agreeing because “[…] it couldn’t get any worse.”, (05). Expectations before surgery were generally modest and survival-oriented, focused primarily on seizure reduction and the possibility of regaining minimal everyday stability.

In contrast, participants with AMD more frequently articulated future-oriented and identity-related hopes (25: “No, I was happy because I was full of hope, like, after this surgery, nothing bad will happen to you anymore.”). Surgery was associated with aspirations of becoming “[…] the old me again, […]” (25), returning to unrestricted driving, resuming professional responsibilities (06: “I just thought, because things were getting more extreme, and I wanted to continue enjoying my job […]”), or relieving family members from the burden of constant vigilance. Unlike the SMD group, fear and uncertainty surrounding surgery were also more openly acknowledged. Overall, the AMD group approached surgery less as a final rescue intervention and more as a possible opportunity for restoration.

### Severe postoperative decline is associated with persistent functional burden

Although both groups reported meaningful postoperative improvements regarding seizure reduction, the SMD group consistently described persistent and functionally disabling cognitive symptoms that remained central to everyday life years after surgery. Participants reported severe word-finding difficulties, inability to retrieve familiar names, impaired new learning, and profound long-term memory problems (05: “Memory lapses, I had them a lot, and I still do.”). One participant described repeatedly returning from the supermarket without essential items despite carrying written lists, requiring multiple trips to complete simple purchases (02). Improvements in seizure frequency were often narrated alongside ongoing cognitive losses. Several participants depicted their postoperative lives as characterized by alternating “good days and bad days” (02), reflecting ongoing instability rather than recovery.

By contrast, postoperative difficulties in the AMD group were generally more circumscribed and less functionally dominant. While memory complaints, fatigue, or temporary speech difficulties were reported, these symptoms were often described as manageable, partially resolving, or limited in impact on occupational and social functioning (“I think a lot of people didn’t really notice the paraphrase, so I just didn’t mention the word. But they only realized it because I started to stutter.”, 09) One participant returned to full-time work within ten months despite residual memory difficulties.

### Work and identity diverge substantially between groups

The clearest distinction between groups concerned occupational functioning and its relationship to identity. In the SMD group, inability to work or fear of losing work capacity became the central theme of postoperative life. Participants described disability pension (05: “I haven’t been working for over a year now - not in a professional capacity - over a year, so almost two years, […] For a year now, I’ve been receiving a full pension.”), prolonged sick leave (03), failed reintegration attempts, and the necessity of concealing cognitive deficits from employers and colleagues. One participant described “having to keep their mouth shut” (03) to avoid losing the position, while another linked the loss of professional competence directly to loss of self-worth and autonomy (02).

These occupational disruptions were closely tied to financial strain and social vulnerability. One participant recounted surviving on less than 1000€ per month while simultaneously paying transportation and medication costs (03). Another participant framed the entire postoperative period around legal and bureaucratic struggles surrounding work incapacity and pension access (01).

In contrast, participants with AMD generally maintained greater continuity in professional and social roles. Several remained employed, returned to work (10: “There weren’t any major changes in my professional life after that, no. I was off work for quite a while, of course.”), or transitioned gradually into retirement. Occupational adaptation was often described as manageable rather than catastrophic (25: “So we complement each other at work. I’m kind of the nurse, and my colleagues are the computer geeks.”). One participant successfully transitioned from full-time work into supported part-time employment before retirement (10).

### Outcome integration and adaptation differ noticeably between groups

Participants with AMD more often described a coherent integration of epilepsy and surgical consequences into an evolving self-concept (09: “You’re no longer the sick person right after surgery. So who am I, then? […] But it’s about exploring yourself again. Getting to know yourself.”). Even when disappointments or emotional sequelae occurred, surgery was predominantly evaluated as beneficial overall (06: “It was just really nice, and the fact that you just didn’t have those fears anymore”). Seizure freedom or seizure reduction remained psychologically central. A minority of participants explicitly described adapting to altered circumstances through psychotherapy, psychosomatic rehabilitation, or gradual lifestyle adjustment (09,10).

In contrast, participants with SMD reported ongoing ambivalence, unresolved grief, and fragmentation of identity (03: “It’s not your fault. You don’t do it on purpose. […] Still, you should always be strong and fight against it. […]. I’m doing just that right now. Just keep going.”). Cognitive impairments were experienced not merely as symptoms but as disruptions of autonomy, competence, and social belonging. Concealment of deficits emerged as a particularly prominent burden, generating chronic psychological strain and social isolation.

Notably, formal psychological or rehabilitative support was largely absent in this group despite substantial functional burden. By contrast, the few participants who received psychotherapy or psychosomatic rehabilitation described these interventions as highly valuable.

## Discussion

Our findings suggest that severe postoperative memory decline should not be viewed simply as the lower end of a continuous distribution of expected cognitive change, but rather as a distinct clinical outcome with disproportionate consequences for patients’ everyday lives. Patients with average versus severe cognitive decline after epilepsy surgery constructed their postoperative experiences in qualitatively different ways. AMD accounts centered around improvement and restoration, with surgery integrated into a continuing sense of self. SMD accounts were more frequently oriented toward persistent symptoms, ongoing treatment, and illness context, such that even meaningful seizure reduction was experienced as secondary to enduring functional difficulties. These patterns were most apparent in the domain of work and identity. Thus, as SMD arises as a rare but underestimated complication of epilepsy surgery, long-term adaptation may depend less on seizure outcome in isolation than on the extent to which cognitive sequelae intrude upon everyday functioning, occupational identity, and the capacity to reconstruct a coherent personal narrative.

A key strength of this study is the use of centre-specific regression based-change norms, allowing identification of patients whose decline exceeds expected longitudinal variation and examination of the subjective meaning of clinically defined decline, addressing a translational gap between neuropsychological outcome classification and patient experience.

The two groups differed not only in outcome but in the expectations through which surgery was evaluated. SMD patients, facing prolonged instability, repeated medication failure, and biographical adversity, approached surgery from therapeutic exhaustion rather than aspiration. Preservation of cognition was rarely an explicit hope, so subsequent severe losses were experienced as unprepared for trade-offs. AMD patients more often held identity-oriented, future-directed expectations and articulated fears beforehand, which allowed even disappointing outcomes such as seizure recurrence to remain narratively interpretable. Hopes appeared to determine which outcome domains became psychologically meaningful in the first place. These observations support an expanded model of postoperative adjustment: established neurological predictors [5,6] remain critical for predicting decline, but seem insufficient to explain how patients integrate it, which additionally reflects biographical resources, illness meaning, and expectation calibration [14,25,26]. Although outcome prediction and the factors underlying these different trajectories were beyond the scope of this study, it is notable that patients with SMD and AMD appeared to differ in their baseline conditions, which may have predisposed them to their respective outcomes. In particular, patients who viewed surgery as a last resort or expressed marked desperation before surgery may warrant closer attention during preoperative decision-making, as these factors could reflect limited psychosocial support, reduced psychological reserve, and lower resilience.

The most clinically important observation concerns the near-complete absence of structured psychological preparation and postoperative rehabilitation in the SMD group. Despite persistent impairments, most participants navigated adaptation alone. Concealment of cognitive deficits was particularly striking: participants actively hid memory and language difficulties from employers and colleagues for fear of stigma and occupational loss, at substantial ongoing psychological cost. Cognitive rehabilitation following TLS has shown promising effects on memory outcomes [28], but the available evidence remains limited, and rehabilitation is not routinely integrated into postoperative care. Likewise, specialized inpatient rehabilitation programs have demonstrated benefits for employment, quality of life, emotional adaptation, and overall health [27–29], yet these interventions have largely been evaluated within research settings rather than implemented as standard clinical care. Recent expert recommendations have emphasized the need for individualized perioperative care, including both prehabilitation and structured postoperative rehabilitation [30]. Our findings complement these recommendations by illustrating the lived consequences when such support is absent.

These findings have direct implications for preoperative care. First, expectation management and structured pre-surgical counselling should be treated as a necessity rather than optional: patients may understand the statistical risk of memory decline without integrating its implications for work, autonomy, and self-concept, so counselling must move beyond narrow informed consent toward explicit expectation calibration [31–33]. Second, the data support structured postoperative pathways combining neuropsychological follow-up with psychological counselling, psychosomatic rehabilitation, and occupational reintegration, targeted especially at patients identified as high-risk through sensitive individual-level metrics [34]. Persistent unemployment and failed reintegration may carry substantial long-term costs, countering this with appropriate investments may improve both patient-centred and health-economic outcomes [27–29].

Several limitations apply: the sample was small and single-centre, the design prioritised experiential depth over representativeness, and retrospective narratives may be colored by the participants’ current status. The AMD comparison group showed some internal heterogeneity, with one case at the 62nd percentile against three clustered in the low 20s. All cases nonetheless met the a priori inclusion criterion, and sensitivity review indicated that this case did not drive the observed between-group differences; however, such within-group variability warrants caution Nevertheless, integrating individually classified objective cognitive outcomes with in-depth narrative analysis offers experiential resolution largely absent from existing literature.

In conclusion, severe cognitive decline was experienced not merely as loss of function but as disruption of identity, autonomy, and anticipated life trajectory. Current pathways insufficiently address these dimensions before and after surgery. Structured expectation management and pre-, peri-, and postoperative counselling as well as rehabilitation therefore appear not merely supportive but potentially essential for helping patients integrate surgical outcomes into sustainable daily lives.

## Contributors

During the preparation of this manuscript, the authors used DeepSeek and ClaudeAI to assist with language editing and refinement of the manuscript text. All AI-generated suggestions were critically reviewed and edited by the authors, who remain fully responsible for the accuracy, integrity, and final content of the manuscript.

## Data availability statement

The qualitative interview data generated and analysed during this study are not publicly available due to the highly confidential nature of the patient interviews and the potential risk of participant identification. Access to the interview guideline and coding scheme is available from the corresponding author upon reasonable request.

## Ethics approval statement

This study was approved by the Ethics Committee of the University Hospital Bonn (approval number: 140/23-EP). All participants provided written informed consent prior to participation. The study was conducted in accordance with the Declaration of Helsinki.

## Contributors

JT and CH had full access to the data in the study and are the guarantors for the integrity of the data and the accuracy of the data analysis. Concept and design: JT, CH, DM, GT, JAW. Acquisition, analysis or interpretation of data: all authors. Drafting of the manuscript: JT, CH, and DM. Critical revision of the manuscript for important intellectual content: all authors. Statistical analysis: JT and DM. Administrative, technical or material support: RS, and CH. Supervision: CH.

